# Hemispheric atrophy as a predictor for naming recovery following left hemisphere ischemic stroke

**DOI:** 10.1101/2025.08.26.25334370

**Authors:** Voss Neal, Andreia V. Faria, Wen Zhang, Argye E. Hillis, Melissa D. Stockbridge

## Abstract

Numerous large-scale epidemiological studies investigating the trajectory of cognitive recovery after ischemic stroke have presented data suggesting an immediate drop in cognition acutely post-stroke followed by persistent, accelerated decline over time when averaged as a group. We sought to further examine this trend, speculating that the average persistent decline may be a reflection of two subgroups with vastly different prognoses: 1) a minority experiencing decline secondary to neurodegenerative processes like vascular dementia and Alzheimer’s disease, and 2) a majority without marked progressive brain atrophy who typically see improvement. Our team thus investigated atrophy’s association with language recovery, hypothesizing that declining naming performance in the year after left hemisphere ischemic stroke would be correlated to atrophy of the contralesional hemisphere. We postulated that volume loss within the lesioned hemisphere would be less informative due to separate confounding processes related to the stroke itself like Wallerian degeneration and encephalomalacia.

Participants (*n*=72; *M*[*SD*] age=60[11]) in a longitudinal cohort study of language following left hemisphere ischemic stroke were included if they completed an MRI both acutely and chronically (either 6- or 12-months post-stroke). Naming performance was assessed using the Boston Naming Test, stroke volumes were extracted from acute imaging, and atrophy was measured as the monthly percent change in hemispheric volume from baseline to chronic scan for each individual. Pearson’s correlations were calculated to determine the relationship between lesion volume and atrophy along with atrophy and change in Boston Naming Test score.

Lesion volume was negatively correlated to the monthly percent change of volume in the left (ipsilesional) hemisphere (r=-0.48; p<0.0001) but was not correlated to rate of right (contralesional) hemisphere volume loss. While there was no clear relationship between atrophy of the left hemisphere and language recovery, we found that volume changes of greater negative magnitude within the right hemisphere (increased atrophy) were associated with worse functional recovery of language (r=0.38; p=0.0025).

By showing that atrophy of the right hemisphere was not significantly impacted by left hemisphere lesion size, we suggest that accelerated volume loss in the non-lesioned hemisphere after stroke may be indicative of a separate pathology. We then go on to support this claim with behavioral data showing that greater rates of volume loss within the non-lesioned hemisphere were associated with poorer naming recovery. Together, these findings imply that contralesional atrophy after stroke may have negative implications for recovery and could serve as a useful imaging signature for separate neurodegenerative processes.

## Introduction

Cognitive impairment^1^ and aphasia^2^ are among the most debilitating sequelae of stroke, posing substantial challenges in patients’ return to work and prior avocations. It is estimated that almost half of left hemisphere stroke survivors deal with some degree of aphasia, ranging from mild anomia to global aphasia.^3^ Cognitive impairment also frequently is reported, affecting as many as 60% of individuals who have had a stroke in either hemisphere.^1^ This can manifest in numerous ways, with some cases of mild cognitive impairment seeing improvement and others progressing to disorders such as vascular dementia and/or Alzheimer’s disease.^1^

The scale of the issue at hand has led numerous institutions to conduct large epidemiological studies aimed at defining the trajectory of cognitive recovery/decline over time.^4-8^ One such longitudinal study by Levine *et al*.^4^ followed 23,572 individuals with or without stroke over a 6-year period and found that the cognitive trajectory of stroke survivors was, on average, characterized by an abrupt, initial decline in cognition followed by a persistent, accelerated decline relative to those who remained stroke-free. This trend has been replicated in other population-based studies,^5-8^ corroborating the notion of an accelerated cognitive decline on average amongst stroke survivors.

This average decline, however, may not reflect cognitive changes in the majority of individuals who have experienced a stroke and instead could be a result of conflating two distinct populations. It is likely that a minority of stroke survivors show marked decline in language while the larger majority sees improvement or complete recovery.^9^ As discussed earlier, the decline experienced by some stroke survivors may be secondary to repeated infarct as well as the development of vascular dementia and/or Alzheimer’s disease, which has been supported by data from both animal and human studies.^10-12^ By one estimate, around one in five patients meet criteria for a diagnosis of vascular dementia within 6 months after stroke.^13^ While the rate of post-stroke incident dementia is significantly greater in cases of repeat stroke,^14^ the threat is still present in the absence of stroke recurrence. A meta-analysis of 73 studies on the emergence of dementia up to 1 year after stroke reported mean prevalence estimates between 7.4-12.0% when examining cases of first-ever stroke specifically.^15^

Previous efforts from our lab have focused on the trajectory of recovery post-stroke as it pertains to language, and the resultant data support our skepticism regarding a central tendency of cognitive decline being ascribed to stroke survivors. In a longitudinal study of the influence of lesion characteristics on changes in picture naming and discourse in the first 6 months after stroke, we found that 71% of 40 people with aphasia showed improvement in picture naming on the Boston Naming Test (BNT), 5% showed no change, and 21% showed decline.^9^

Despite a great deal of relevant data from extensive epidemiological studies, it is still largely unclear as to why some individuals (15-25%) show persistent decline, rather than recovery, after stroke. Some scholars posit that atrophy may be a driver for the cognitive decline seen after stroke and that associated imaging signatures could be clinically informative.^16-18^ Data from a multicenter, observational cohort in Australia showed that change in total brain volume was greater in stroke survivors who were cognitively impaired than those without deficits^16^ and that brain volume loss was greater ipsilesionally than contralesionally when examining both groups as a whole.^19^ Interestingly, further analysis showed that cortical thinning was greater in the contralesional cortex of cognitively impaired stroke survivors than in the ipsilesional cortex of cognitively normal stroke survivors,^20^ suggesting that the contralesional hemisphere may be informative in predicting cognitive decline.

Here, we further investigate the impact of atrophy on language recovery or decline following ischemic stroke. Our aim is to elucidate neural and functional differences present among stroke survivors that could explain why some experience decline and others experience improvement. Thus, we hypothesized that decline in language and cognitive function in the year after left hemisphere ischemic stroke correlates with degree of atrophy in the contralesional hemisphere. We focus on atrophy in the contralesional hemisphere because atrophy in the lesioned hemisphere includes Wallerian degeneration and encephalomalacia due to the stroke itself. Confirmation of our hypothesis would provide evidence that cognitive decline after stroke is due to development or unmasking of neurodegenerative disease which may or may not be independent of the stroke.

## Materials and methods

### Participants

We analysed data collected from 72 participants (34 female; *M*[*SD*] age=60[11]) as part of an ongoing longitudinal cohort study following patients’ recovery during the year after acute left-hemisphere ischemic stroke. This study includes a battery of multiple behavioral assessments primarily focusing on language and cognitive performance in conjunction with a neuroimaging component. Participants were selected from this study for our analysis if they had completed an MRI acutely in addition to a follow-up MRI at either 6-or 12-months, utilizing the more chronic of the two if both follow-ups were completed. Data collection for the resultant sample occurred between 2008 and 2024. The records reviewed were an exhaustive set who met inclusion criteria and had MRI imaging data at the required timepoints, leading to our study size of 72. We examined participants’ behavioral data along with the imaging analysis, focusing on the BNT (30-item edition) at the timepoints with completed MRIs as a proxy for naming and language function. Participants (or their legally authorized representatives in cases of severe cognitive impairment resulting from the stroke) were approached if they had experienced a left hemisphere ischemic stroke outside of the brainstem/cerebellum, were over the age of 18, were proficient English speakers, and had no history of hearing/vision impairment. Study team members screened medical charts to identify and exclude patients with prior neurological disorder including, but not limited to, dementia and mild cognitive impairment.

We sought to eliminate collection bias within the population by approaching all patients meeting eligibility criteria. Those who consented to participate then completed acute testing within 5 days of symptom onset. In cases where the patient was intubated over the course of their hospitalization, the acute window was extended to include the 5 days post-extubation. Following discharge, members of the study team reached out to patients and/or family members via phone call to schedule follow-up visits at 3-, 6- and 12-months after hospitalization, though data collected at 3-month follow-ups was not included in this analysis. To minimize loss to follow-up, multiple contact attempts were made per timepoint window along with offering transportation assistance within reasonable distance. Participants were encouraged to complete all clinically indicated rehabilitation including speech language pathology to address deficits in communication and were not barred from participating in other interventional or longitudinal research that might influence their results. The study was conducted at Johns Hopkins Hospital and Johns Hopkins Bayview Medical Center in accordance with the Johns Hopkins Medicine Institutional Review Board (IRB Protocol NA00042097), and participant consent was obtained according to the Declaration of Helsinki.

### Naming assessment

Participants completed a 30-item version of the Boston Naming Test (BNT)^21,22^ during the acute phase of their recovery and again at either 6- or 12-months post-symptom onset, using the more chronic scores for analysis. Administrators of the BNT presented an illustrated list of 30 common objects to name. This metric is particularly sensitive for detecting mild anomia, a common residual deficit in persons with aphasia.^23^ Of the 72 participants with acute and chronic imaging data, 62 had associated BNT scores at both timepoints. The 10 who did not have available data at either one or both timepoints were thus unable to yield a change in BNT score and were not included when analyzing the correlation between atrophy and naming recovery but were included in our analysis regarding the correlation between lesion size and atrophy. Missing data for these participants were a result of the inability to complete acute testing prior to hospital discharge, omission of the assessment at follow-up, or the completion of testing during a brief window when the BNT was not included in the behavioral protocol.

### Healthy Controls

As part of a preliminary analysis intended to contextualize the atrophy seen in our population of stroke survivors, we also examined rates of volume loss associated with healthy aging for neurotypical controls (n=11) who had an initial MRI followed by a subsequent scan 1 year later. All controls were without diagnosis of any neurological disease and had Montreal Cognitive Assessment (MOCA) scores >26.

### Imaging analysis

#### Acquisition and processing

The baseline image is the scan completed at the time of admission for first symptomatic stroke on a 3 Tesla scanner. All the patients had diffusion-weighted imaging (DWI), which was used to delineate the stroke core and obtain its volume. This was done with Acute-stroke Detection Segmentation (ADS), an automated tool for ischemic stroke quantification.^24^ A subset of 43 patients had high resolution T1-weighted imaging (T1-WI), specifically Magnetization-Prepared Rapid Gradient Echo (MPRAGE), at baseline; 29 had regular T1-WI with slice thickness varying from 4 to 6mm. All the patients had T1-WI (MPRAGE) at follow-up, with 56 patients having scans available from their 1-year follow-up and 16 patients having scans available from their 6-month follow-up. The 16 patients without 12-month follow-up imaging data were either lost to follow-up after numerous contact attempts or declined an MRI at the final timepoint, as imaging was offered as a supplemental portion of the longitudinal protocol related to language recovery. We addressed the discrepancy that arose from grouping 6- and 12-month scans by calculating the rate of atrophy as a monthly % change, which is described in further detail later on. Follow-up scans were performed in the F.M. Kirby Research Center at Kennedy Krieger Institute and completed at the same time point as the BNT. The images were segmented using SynthSeg^25^; in this study, we analysed the hemisphere volumes (Fig. 1). These volumes were calculated on synthetic MPRAGEs generated for each case with SynthSR^26^ to minimize the differences in the image protocols. Quality control for all the steps was done by a radiologist. Image analysis for the controls was done at baseline and again after 6- and 12-months using T1-WI MPRAGE in an identical post-processing pipeline as used for the patients.

**Figure 1.**
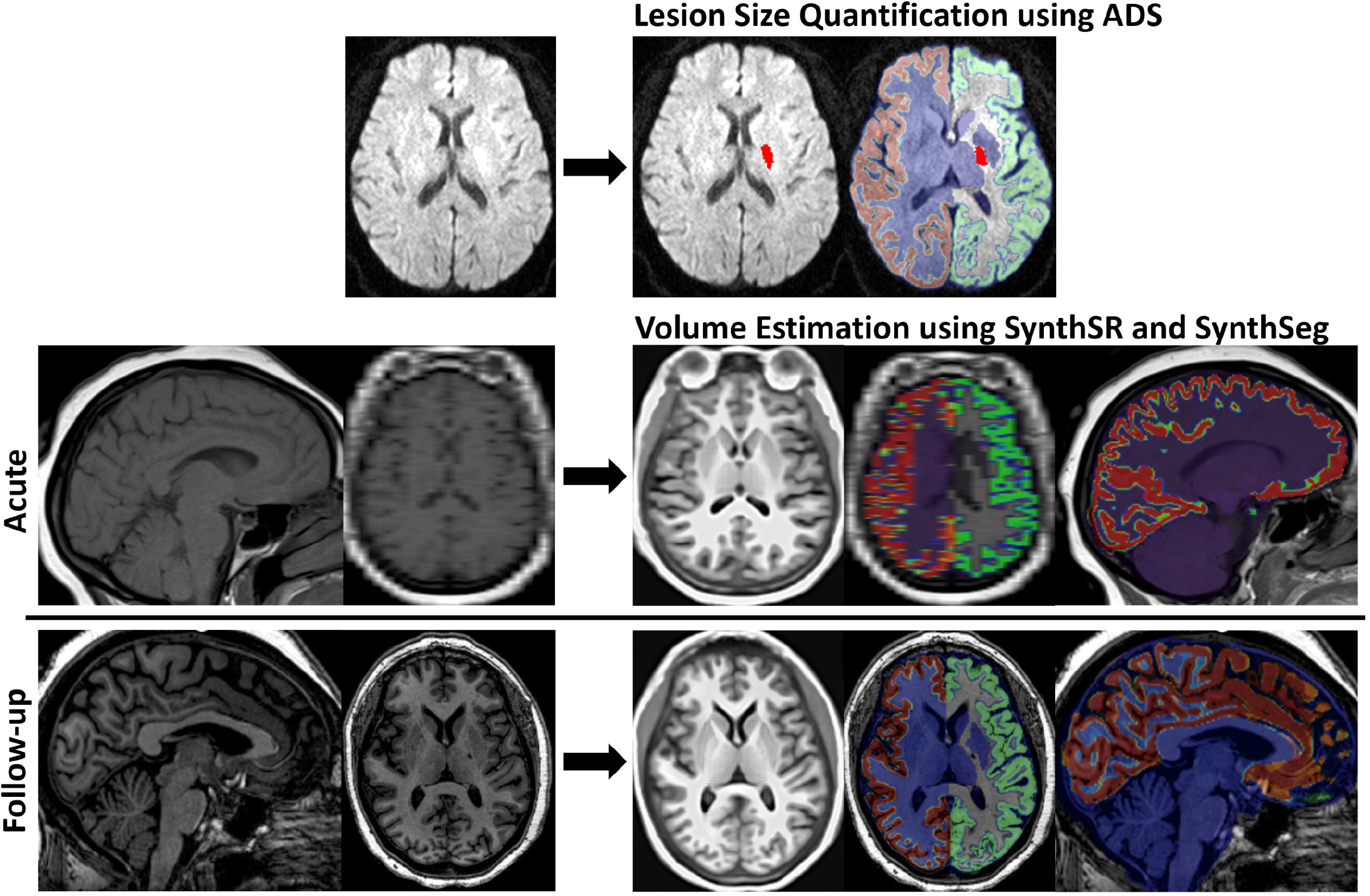
Schematic representation of the imaging analysis. Lesion size was extracted from acute Diffusion Weighted Imaging (DWI) scans using the Acute stroke Detection and Segmentation (ADS)^24^ tool, as denoted by red overlay. Synthetic Magnetization-Prepared Rapid Gradient Echo (MPRAGE) scans were created from T1-weighted images using SynthSR^26^ and subsequently segmented using SynthSeg^25^ to obtain volumes of diverse brain regions (depicted by the colored overlay). In this study, we analyzed hemisphere volumes only, obtaining measurements acutely and at chronic follow-up to quantify atrophy.

#### Atrophy quantification

Atrophy was measured by comparing the volumes of the hemispheres at baseline scans to follow-up scans for each individual, as (follow-up – baseline)/baseline. This allowed us to minimize variation between head volumes and normalize the range of values. Furthermore, we divided this ratio by the number of months between the time points. In summary, the resultant index is a % change from the initial volume, per month, with negative numbers representing atrophy (i.e., follow-up<baseline). In order to make the volumetric changes comparable to the naming changes, changes in naming scores were also divided by the time interval between them, representing the monthly rate as well.

#### Statistical analysis

Group differences in structural volumes (e.g., ventricles, hemispheres) were assessed using *t*- tests. Pearson’s correlation was used to assess relationships between atrophy, BNTs, and stroke volumes. Statistical tests were two-tailed and utilized a significance level of α=0.05 to denote the probability of a Type I error. Missing data were not imputed prior to analysis.

## Results

We reviewed 732 records of consented participants in our longitudinal study of language recovery following left hemisphere ischemic stroke. Of this group, 72 participants were included in this sample having met eligibility criteria with sufficient MRI imaging at both the acute and chronic timepoints. Table 1 shows the profile of the 72 participants along with summary statistics regarding lesion volume, naming performance, and atrophy rates for both hemispheres. Controls used in our preliminary analysis are also included and had age and gender distribution not significantly different from the participants.

**Table 1.** Demographic and Summary Statistics.

|  | Stroke Survivors (*n* = 72) |  | Controls (*n* = 11) |  |
| --- | --- | --- | --- | --- |
|  | *M* (*SD*) | Range | *M* (*SD*) | Range |
| Age (years) | 60 (11) | 28 to 85 | 56 (15) | 39 to 77 |
| Education (years) | 14 (3) | 9 to 20 | - | - |
| Lesion Volume (cc) | 18.89 (33.64) | 0.20 to 181.07 | - | - |
| Naming Performance (*n* = 62) |  |  |  |  |
| Acute BNT Score (/30) | 20 (9) | 0 to 30 | - | - |
| Chronic BNT Score (/30) | 24 (7) | 0 to 30 | - | - |
| Change in BNT Score | 4 (6) | -5 to 26 | - | - |
| Atrophy |  |  |  |  |
| Change in LH Volume (%) | -3.49% | -11.01% to 0.74% | -0.85% | -1.71% to -0.11% |
| Change in RH Volume (%) | -1.84% | -7.21% to 3.16% | -0.84% | -1.77% to 0.08% |
| Sex (Female:Male) |  | 38:34 |  | 3:8 |
| Race (Black:White:Other) |  | 44:27:1 |  | 0:11:0 |
| Ethnicity (Non-Hispanic:Hispanic) |  | 71:1 |  | - |
| Handedness (Left:Right) |  | 9:63 |  | 0:11 |

BNT = *Boston Naming Test – Shortened* (Kaplan, 2016; Mack, 1992); LH = Left hemisphere; RH = Right hemisphere.

### Atrophy associated with healthy aging compared to post-stroke

Both controls and patients showed reduction in the brain volume over time, as well as ventricle enlargement, as expected with aging. However, the atrophy was more severe in the patients than in controls (p=0.028, effect size by Cohen’s d=0.545, power=0.80; Supplementary Fig. 1).

### Lesion volume and subsequent hemispheric atrophy

While volumetric reduction in the left hemisphere correlated to stroke volumes (r=-0.48; p<0.0001; Fig. 2A), indicating that it might be the direct result of the stroke as well as secondary Wallerian degeneration, atrophy of the right hemisphere did not (Fig. 2B), suggesting a minimal direct effect of the stroke on progressive atrophy in the non-lesioned hemisphere.

**Figure 2.**
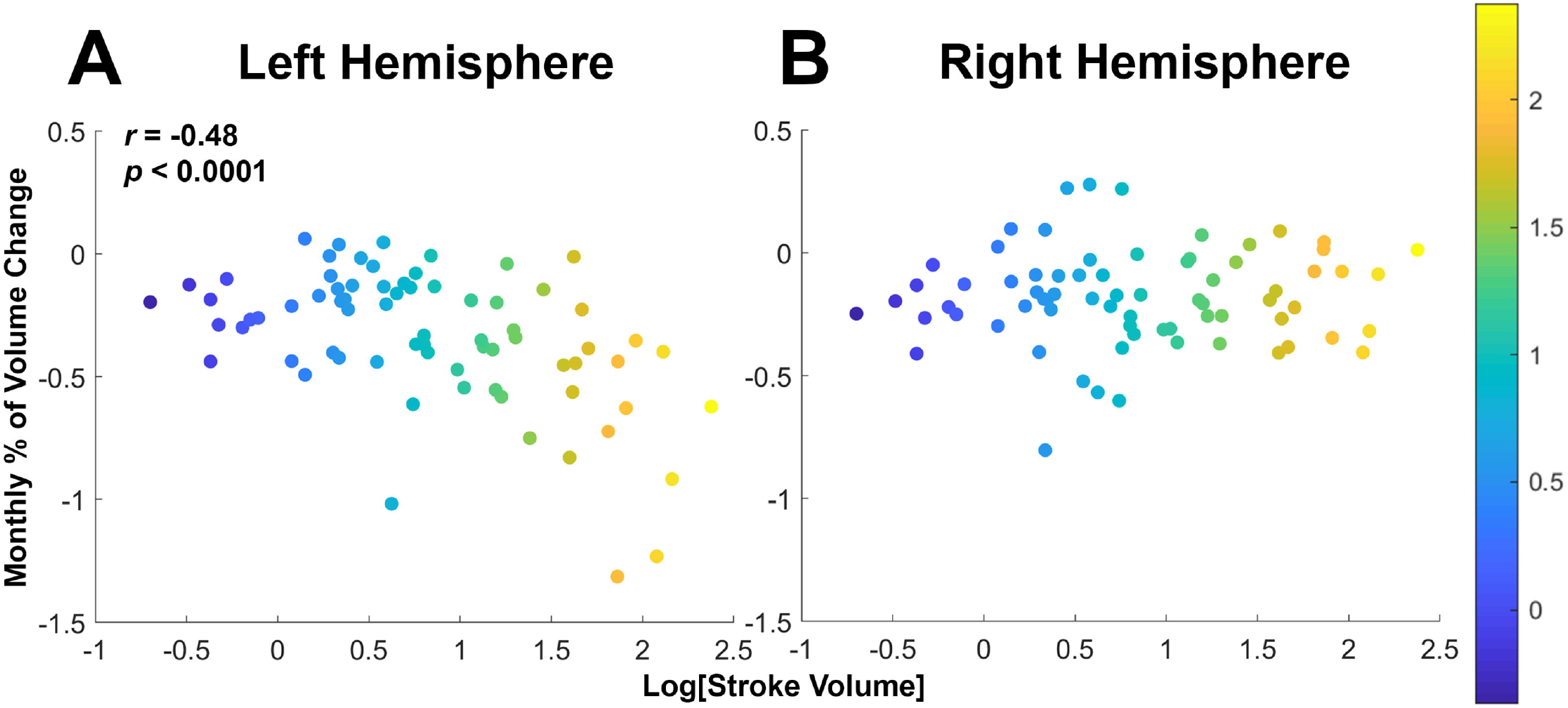
Scatter plots of stroke volume versus hemispheric atrophy. Stroke volume correlated to atrophy of the left hemisphere (**A**), but not of the right hemisphere (**B**). The color scale shows Log[Stroke Volume] in mL.

### Lesion volume and naming recovery

When examining the relationship between stroke volume and BNT scores, an interesting phenomenon emerges. Patients with larger strokes appear to recover more (r=0.41; p=0.001; Fig. 3A). However, this is largely due to a ‘ceiling effect’—patients with high baseline scores (BNT>25) have less room for improvement. Once these patients are excluded, no clear relationship between stroke volume and recovery is observed (Fig. 3B).

**Figure 3.**
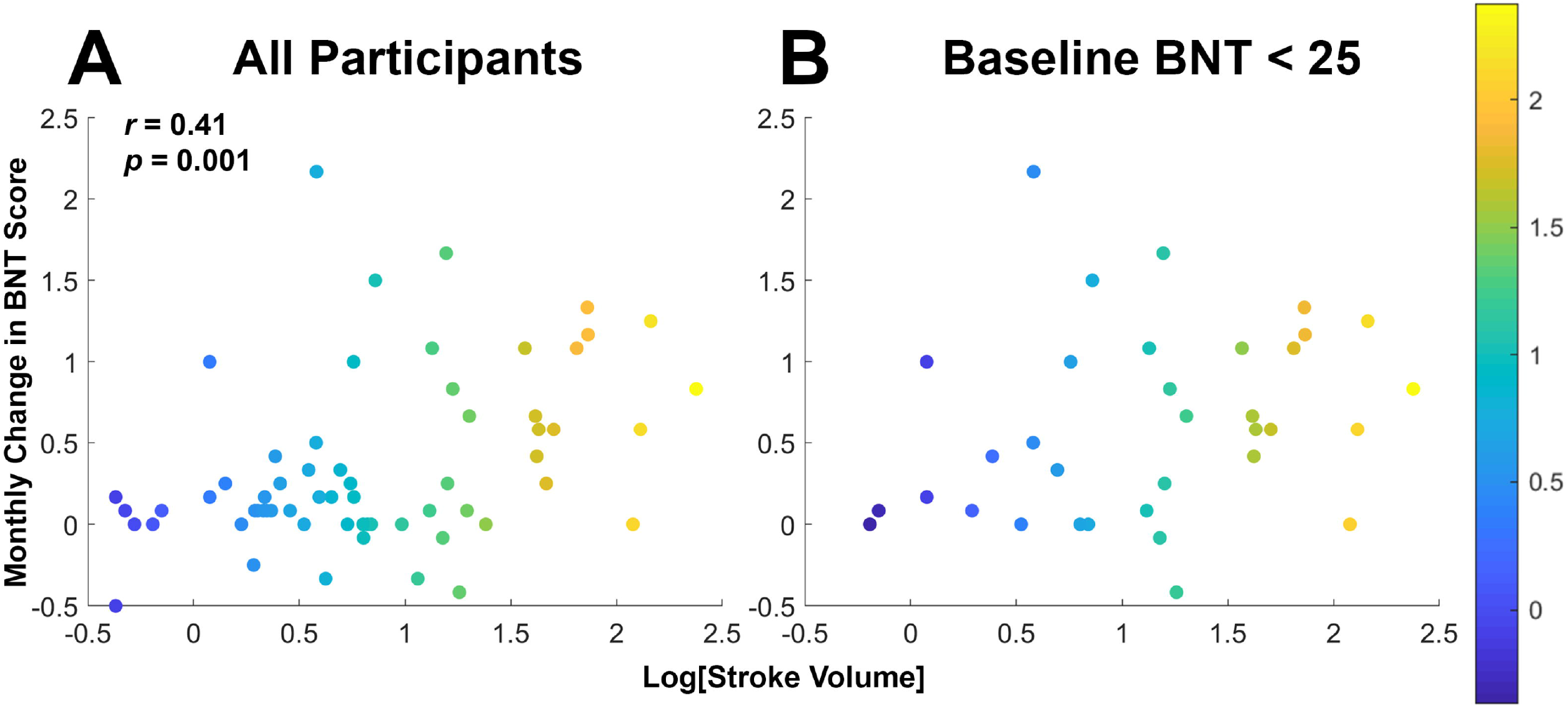
Scatter plots of stroke volume versus change in naming. Patients with large strokes counterintuitively appear to recover more (**A**), which can be explained by “ceiling effects.” When patients close to naming ceiling at baseline are removed – those with less room to recover – this relation disappears (**B**). The color scale shows Log[Stroke Volume] in mL.

### Hemispheric atrophy and naming recovery

The relationship between language recovery and atrophy in the left hemisphere was not very informative, as its volume is heavily influenced by stroke volume, as previously noted. In fact, a similar counterintuitive pattern is observed: patients with greater left hemisphere atrophy (which we saw was correlated to larger strokes) appear to recover more (Fig. 4A). Again, this phenomenon could be explained by a ceiling effect and disappears when excluding patients with high naming score at baseline. In contrast, in the right hemisphere— where atrophy is not influenced by stroke volume—there is a clearer relationship: patients with greater atrophy tend to show less functional recovery (r=0.38; p=0.0025; Fig. 4B). This relation persists when excluding patients with high naming scores (r=0.36; p=0.039; Supplementary Fig. 2A) or smaller strokes (r=0.48; p=0.0014; Supplementary Fig. 2B) at baseline.

**Figure 4.**
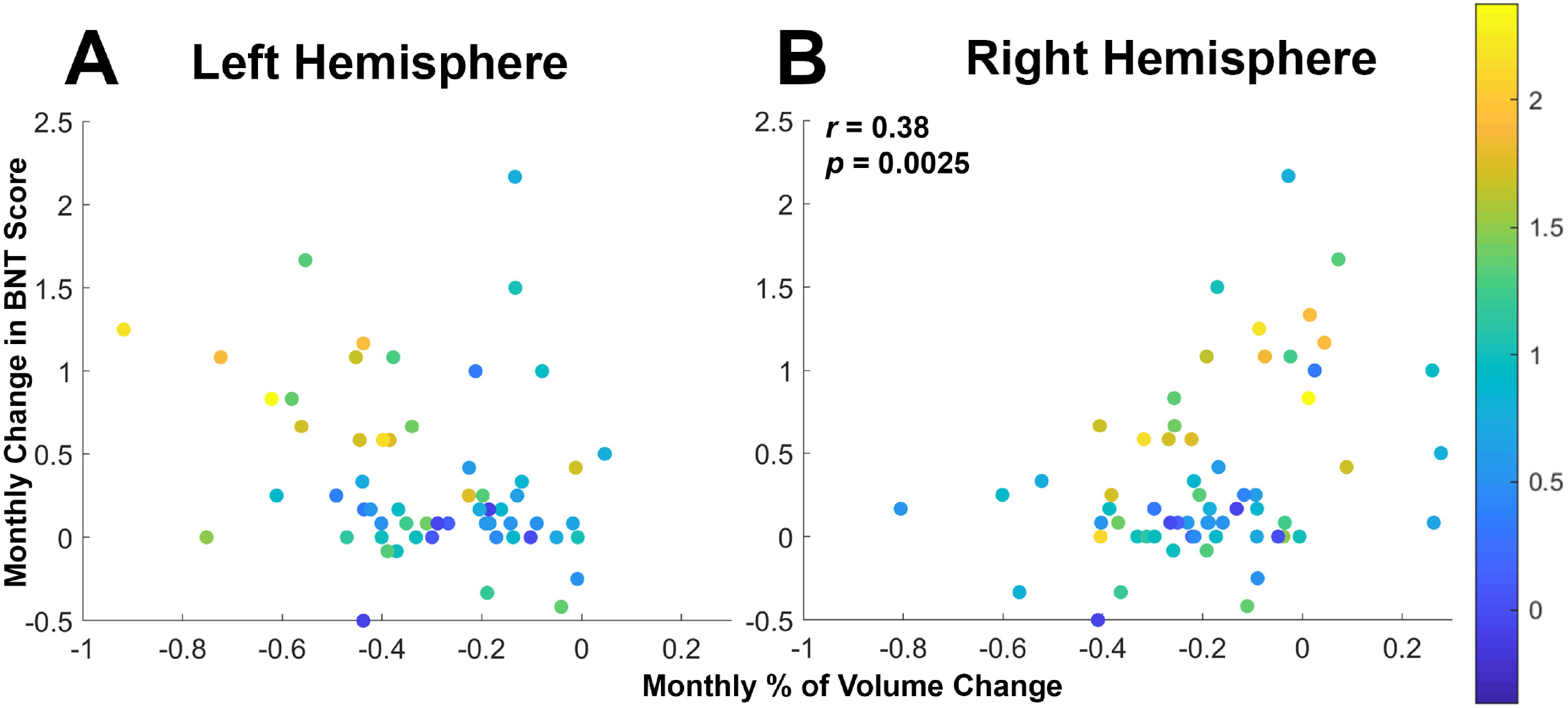
Scatter plots of hemispheric atrophy versus change in naming. Atrophy of the left hemisphere, highly influenced by the stroke volume, was not correlated to naming recovery (**A**), while atrophy of the right hemisphere was associated with worse recovery of naming (**B**). The color scale shows Log[Stroke Volume] in mL.

## Discussion

Given the growing societal impact of cognitive decline following stroke and the importance of better understanding the trajectory of recovery for stroke survivors, we set out to examine the role that atrophy plays in post-stroke cognitive impairment and aphasia. Specifically, our team used longitudinal behavioral and imaging data to characterize the effect of left hemisphere ischemic stroke and lesion volume on rates of hemispheric atrophy in the year after stroke and then further explored the significance of atrophy in relation to naming recovery, as measured by performance on the Boston Naming Test.

As expected, participants with larger strokes saw greater rates of atrophy of the left, lesioned hemisphere (likely due to Wallerian degeneration and encephalomalacia), but we found no significant relation between lesion volume and atrophy of the right, contralesional hemisphere. Thus, we surmised that accelerated rates of contralesional atrophy in stroke survivors may indicate a separate neurodegenerative process, such as vascular dementia.

To expand upon that observation, we investigated contralesional atrophy as a potential imaging signature for cognitive decline and sought to determine if volumetric reductions in the non-lesioned hemisphere manifested behaviorally as a predictor of naming recovery or decline. Results aligned with our hypothesis, showing that participants with greater rates of atrophy in the right hemisphere exhibited worse functional recovery of naming. In contrast, patients showing minimal right hemisphere atrophy tended to see improvement, rather than continued decline, in naming over the first year after stroke.

Involvement of the contralesional hemisphere in the recovery process after stroke has been documented, with prior research indicating that increased contralesional activity, as a compensatory mechanism, contributes to motor recovery following stroke^27^ and may be increasingly important for patients with larger lesions.^28^ Heiss^29^ drew similar conclusions with respect to aphasia, stating that although the greatest potential for recovery comes from reorganization of the ipsilesional hemisphere, compensatory utilization of the contralesional hemisphere can still lead to improvement (albeit to a lesser degree) in cases of larger lesions where the language network is sufficiently compromised. The higher utility of maintaining left hemisphere functionality as opposed to right hemisphere compensation is reflected with work by Keser *et al*.,^30^ who utilized diffusion tensor imaging (DTI) acutely and chronically along with the BNT in left hemisphere stroke survivors. They concluded that greater recruitment of the right hemisphere’s language homologues was actually associated with poorer language outcomes, presumably as it reflects difficulty with utilization of the dominant hemisphere.

Our finding that left hemisphere stroke volume was unrelated to right hemisphere atrophy indicates that accelerated contralesional atrophy may be a harbinger for separate neurodegenerative processes leading to the decline. Although the development of contralesional hemisphere atrophy may be causally related to stroke, when it is due to vascular dementia or Alzheimer’s dementia, it is not an inevitable consequence. Future studies are essential (and many are ongoing) to identify why some individuals develop dementia after stroke.

Previous literature examining the longitudinal impact of stroke on cognition provides invaluable insight regarding the average trajectory of recovery for stroke survivors, largely analyzing their samples as a whole. Here we propose a further granulation of this population along the lines of contralesional atrophy to help identify groups within the population that may experience vastly different trajectories of recovery. Our team contributes a novel lens to the literature by pairing longitudinal MRI imaging with behavioral data to examine the correlation between atrophy of the right hemisphere and naming accuracy during the year after left hemisphere ischemic stroke. Through this lens, we hope to partially address the uncertainty regarding why some individuals experience continued decline after stroke and others see improvement.

These results shine light on the potential neurodegenerative implications of post-stroke contralesional atrophy and its downstream effect on language recovery. While this has merit on its own, our research may provide greater predictive value if considered as a singular component for future multifactorial models. Lesion characteristics like infarct size and location,^31^ overall brain health based on factors such as white matter hyperintensities^32,33^ and brain age,^34^ and other predictive factors including demographics^35^ and social determinants of health^36^ are all potential contributors to future models, though this list is by no means exhaustive.

Better predictive tools in the hands of clinicians would in turn improve the efficacy of targeted therapies and lifestyle modification to optimize patient outcomes and help maintain function. Intervention could take on various forms, with the most established practices promoting vascular risk management^37^ and cognitive engagement.^38^ Subsequent studies will hopefully support additional methods for improving patient outcomes post-stroke when paired with more accurate means of prediction. Areas of interest include, but are not limited to, anti-inflammatory therapies,^39^ gene-based approaches,^40^ and non-invasive stimulation techniques leveraging the brain’s neuroplastic potential, such as Transcranial Magnetic Stimulation ^41^ and Transcranial Direct Current Stimulation.^42^

The long-term ramifications of our findings are limited by the time frame with which data collection occurred, as no follow-ups were completed beyond the 1-year mark, nor did we have any data prior to participants’ hospital admission to determine cognitive trajectory before their stroke. Other larger scale epidemiological studies of post-stroke cognitive decline have followed their cohort for years before and after stroke, though typically with a trade-off in the ability to attain repeated imaging. Our findings would be strengthened if follow-up data were available at time points beyond the 1-year mark, and future work may be beneficial to determine how the trends presented here hold up when assessed further along the course of stroke recovery. Results from the ongoing DISCOVERY study (NCT04916210) will likely provide insights.

There are limitations to our study. We must be careful not to overstate the generalisability of our findings to cognitive decline as a whole, as the correlation presented is between hemispheric atrophy and naming performance within left hemisphere ischemic strokes specifically. There are certainly other domains across which language may be impacted, and still more domains across which cognition in general may be impacted after stroke. Therefore, it will be valuable going forward to examine if greater contralesional atrophy is similarly correlated to decline in other areas such as memory and executive function. It will also be pertinent to examine if this trend persists when hemispheres are reversed (in cases of right hemisphere lesions), a distinction that is not easily examined with language due to its mainly left hemisphere-dominant nature. Finally, plasma or cerebrospinal fluid biomarkers of specific dementias, such as Alzheimer’s disease, would be useful to confirm the presence of independent neurodegenerative diseases.

In conclusion, we utilized longitudinal and imaging data collected both acutely and chronically (either 6-or 12-months) from 72 individuals after left hemisphere ischemic stroke to investigate the relationships between 1) stroke size and hemispheric atrophy and 2) hemispheric atrophy and recovery of naming performance. Our analysis revealed a significant correlation between stroke volume and the rate of atrophy in the lesioned (left) hemisphere, but not of the contralesional (right) hemisphere, implying that accelerated atrophy of the noninfarcted hemisphere may indicate separate pathologies. We then saw that atrophy of the contralesional hemisphere had a significant negative impact on change in naming accuracy, but the same negative impact was not correlated with atrophy of the lesioned hemisphere. Combining these two findings led us to conclude that atrophy of the contralesional hemisphere may have negative implications for language recovery and could serve as a potential imaging signature for the onset of post-stroke cognitive decline caused by neurodegenerative processes such as vascular dementia and/or Alzheimer’s disease.

## Data availability

These data are available from corresponding author following written request resulting in a formal data sharing agreement with Johns Hopkins University School of Medicine. Data from clinical patients are not publicly available in a repository, as consent forms predated current sharing requirements, and it was not practicable to recontact patients for reconsent.

