## Supplementary Figure 1 for "Hemispheric atrophy as a predictor for naming recovery following left hemisphere ischemic stroke"

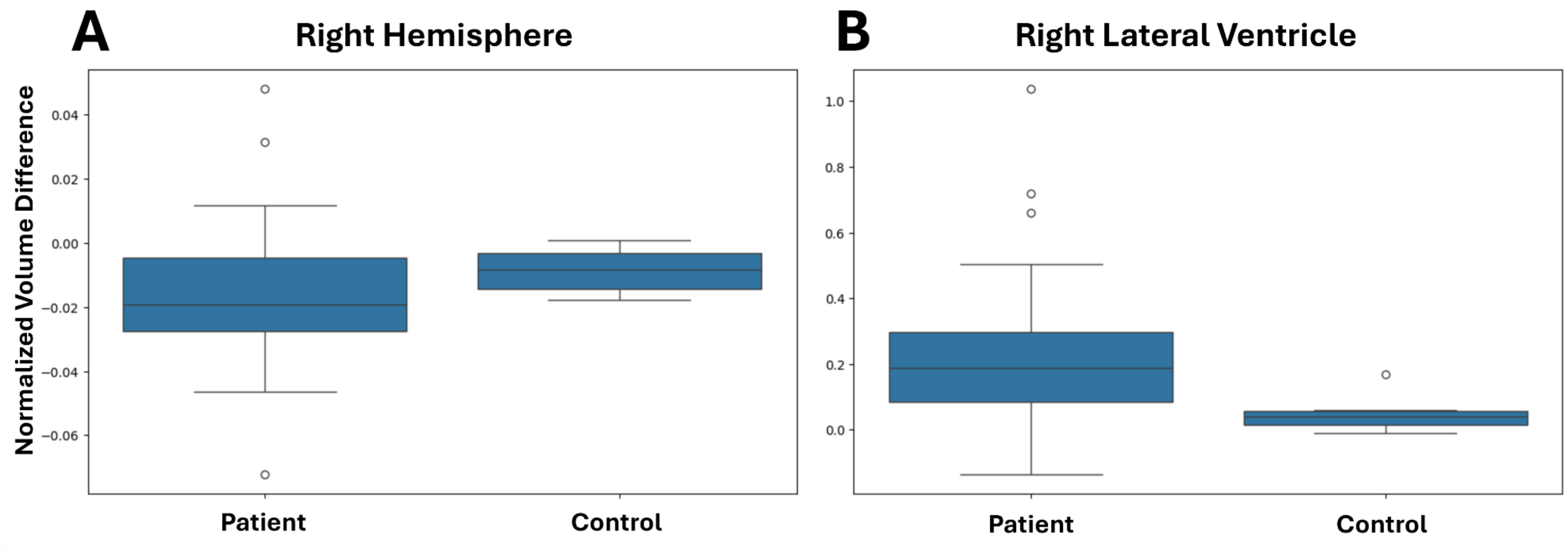


**Supplementary Figure 1 Longitudinal changes in brain volumes**. There is a tendency for right hemispheric volume to decline (**A**) and ventricular volume to increase (**B**) in both controls and patients, but the change in volumes in patients is greater and more variable.
