## Supplementary Figure 2 for "Hemispheric atrophy as a predictor for naming recovery following left hemisphere ischemic stroke"

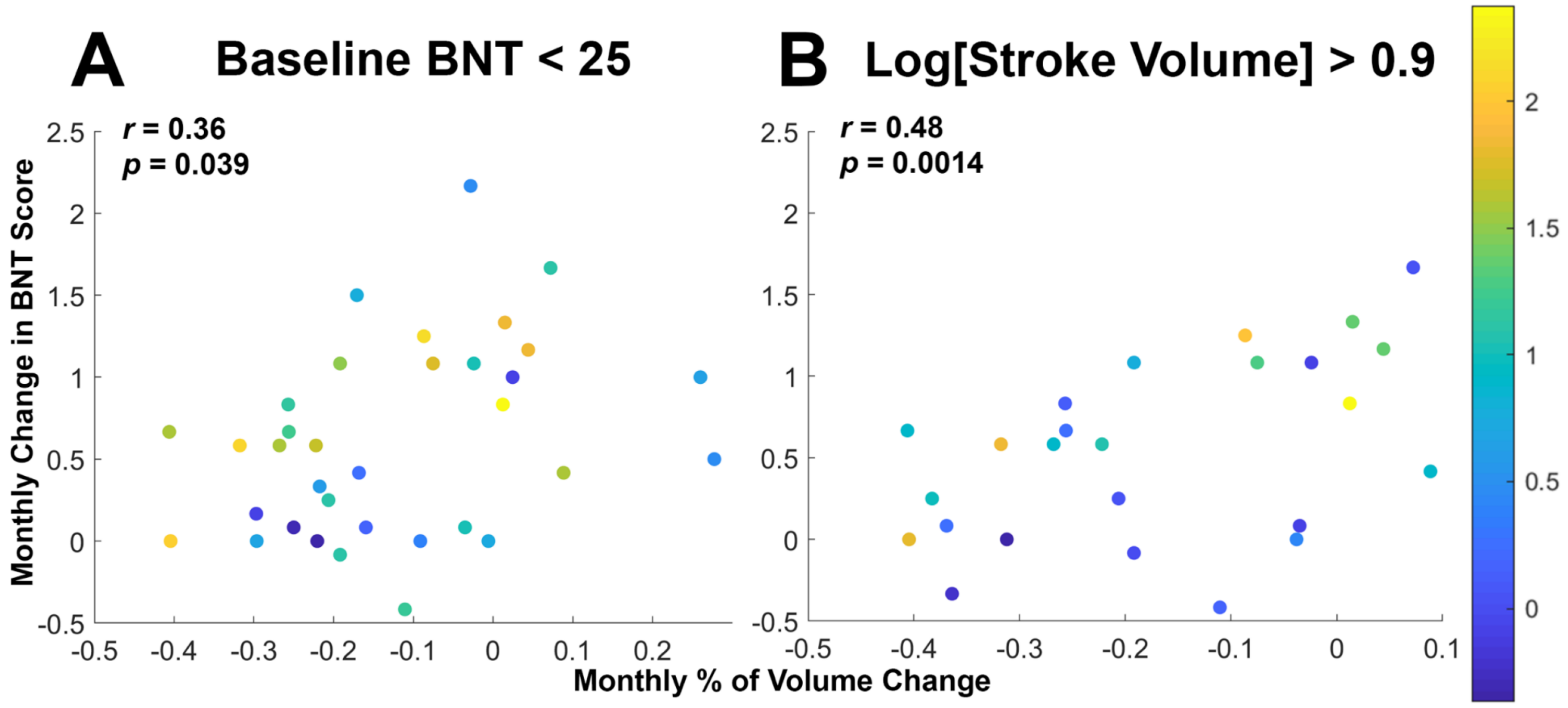


**Supplementary Figure 2 Scatter plots of right hemisphere atrophy versus change in naming.** Patients with greater atrophy tend to show less recovery, a trend that is maintained when excluding both high performers at baseline (**A**) and those with smaller strokes (**B**). The color scale shows Log[Stroke Volume] in mL.
